# Extracorporeal membrane oxygenation in COVID-19 patients and in-hospital mortality: results from the Brazilian Registry using a propensity score matched analysis

**DOI:** 10.1101/2021.05.05.21256475

**Authors:** Daniela Ponce, Milena Soriano Marcolino, Magda Carvalho Pires, Rafael Lima Rodrigues de Carvalho, Heloisa Reniers Vianna, Matheus Carvalho Alves Nogueira, Fernando Antonio Botoni, Fernando Graça Aranha, André Soares de Moura Costa, Giovanna Grunewald Vietta, Felipe Ferraz Martins Graça Aranha, Maria Clara Pontello Barbosa Lima, Ana Paula Beck da Silva Etges, Antonio Tolentino Nogueira de Sá, Luana Martins Oliveira, Carisi Anne Polanczyk

**Author notes:** These authors contributed equally to this work. (Ponce D -). (Marcolino MS -). (Pires MC –). (Carvalho RLR -). (Vianna HR -). (Nogueira MCA -). (Botoni FA -). (Aranha FG -). (Costa ASM -). (Vietta GG -). (Aranha FFMG -). (Lima MCPB -). (Etges APBS -). (Sá ATN -). (Oliveira LM -). (Polanczyk CA -).

## Abstract

Around 5% of coronavirus disease 2019 (COVID-19) patients develop critical disease, with severe pneumonia and acute respiratory distress syndrome (ARDS). In these cases, extracorporeal membrane oxygenation (ECMO) may be considered when conventional therapy fails. This study aimed to assess the clinical characteristics and in-hospital outcomes of COVID-19 patients with ARDS refractory to standard lung-protective ventilation and pronation treated with ECMO support and to compare them to patients who did not receive ECMO. Patients were selected from the Brazilian COVID-19 Registry. At the moment of the analysis, 7,646 patients were introduced in the registry, eight of those received ECMO support (0.1%). The convenience sample of patients submitted to ECMO was compared to control patients selected by genetic matching for gender, age, comorbidities, pronation, ARDS and hospital, in a 5:1 ratio. From the 48 patients included in the study, eight received ECMO and 40 were matched controls. There were no significant differences in demographic, clinical and laboratory characteristics. Mortality was higher in the ECMO group (n = 7; 87.5%) when compared with controls (n = 17; 42.5%), (p=0.048). In conclusion, COVID 19 patients with ARDS refractory to conventional therapy who received ECMO support had worse outcomes to patients who did not receive ECMO. Our findings are not different from previous studies including a small number of patients, however there is a huge difference from Extracorporeal Life Support Organization results, which encourages us to keep looking for our best excellence.

## Introduction

Despite the fact that the majority of coronavirus disease 2019 (COVID-19) patients develop mild to moderate symptoms, around 5% of people have critical disease, with severe pneumonia and acute respiratory distress syndrome (ARDS)[1]. In these cases, therapy includes corticosteroids, protective pulmonary mechanical ventilation, neuromuscular blockade, higher positive end-expiratory pressure, pulmonary recruitment techniques, prone positioning, and more recently tocilizumab[2]. For refractory cases, extracorporeal membrane oxygenation (ECMO) may be considered a last resort in certain patients with critical pulmonary compromise[3]. Previous studies reported success with ECMO in critically ill patients diagnosed with Middle Eastern respiratory syndrome (MERS) and this evidence encouraged physicians to try ECMO in COVID-19 patients presenting ARDS refractory to standard lung-protective ventilation and pronation[3, 4].

In the largest randomised controlled trial on ECMO for ARDS, the ECMO to Rescue Lung Injury in Severe ARDS (EOLIA) trial, there was no difference in 60-day mortality between the ECMO group (35%) and the conventional group (46%); RR 0.76, 95% CI 0.55–1.04; p=0.09)[4]. A post-hoc analysis of the EOLIA trial[4], a meta-analysis of trials of ECMO for ARDS in adults[5], and a network meta-analysis[6] provided credible support to the existence of a survival benefit of ECMO in refractory ARDS.

Despite such optimism for a possible role for ECMO in those cases, early reports of patients with COVID-19 requiring ECMO suggested that mortality could be greater than 90%[7-10]. The Extracorporeal Life Support Organization (ELSO) has been conducting real-time tracking of all COVID-19 cases who received ECMO worldwide, and current results show an estimated cumulative incidence of in-hospital mortality 90 days after the initiation of ECMO of 50% worldwide, and 4% of the patients who did not die are still in the intensive care unit, 11% were discharged to another hospital and 13% were discharged alive to long-term acute care or unspecified location[11]. These findings are less enthusiastic than previous results from a cohort study published by the same group, and there is currently insufficient high quality evidence to recommend either for or against ECMO in patients with COVID-19[12].

Given the lack of clinical trials and prospective studies on ECMO support in COVID 19 patients, questions regarding its effectiveness and feasibility in clinical practice remain unknown. Therefore, we aimed to assess the clinical characteristics and in-hospital outcomes of COVID-19 patients with ARDS refractory to standard lung-protective ventilation and pronation treated with ECMO support and to compare them to patients who did not receive ECMO.

### Methodology

This manuscript adheres to the Strengthening the Reporting of Observational Studies in Epidemiology (STROBE) guideline[13].

Patients were selected from the Brazilian COVID-19 Registry, a prospective multicenter cohort conducted in 37 Brazilian hospitals located in 17 cities from five Brazilian states (Minas Gerais, Pernambuco, Rio Grande do Sul, Santa Catarina, São Paulo). Patients were admitted from March 1 to September 30, 2020. COVID-19 diagnosis was laboratory-confirmed, according to the World Health Organization guidance[14]. Details of the cohort were published elsewhere[15, 16].

For the present study, COVID-19 patients with ARDS refractory to standard lung-protective ventilation and pronation and that received ECMO were selected.

Indications for ECMO support were at discretion of attending teams, according to local protocols (Table 1). Public hospitals do not offer ECMO support in Brazil. In all of them, the costs were paid by the family, and at that time there was no policy of reimbursement of ECMO costs by health insurances in Brazil.

**Table 1.** Indications for extracorporeal membrane oxygenation.

| **Centers** | **Indications** |
| --- | --- |
| Hospital A | PaO<sub>2</sub>/FiO<sub>2</sub> ratio <80 mmHg for more than 6 hours, PaO<sub>2</sub>/FiO<sub>2</sub> ratio <60 mmHg for more than 3 hours or bronchopleural fistula |
| Hospital B | PaO<sub>2</sub>/FiO<sub>2</sub> ratio <80 mmHg for more than 6 hours, PaO<sub>2</sub>/FiO<sub>2</sub> ratio <60 mmHg for more than 3 hours or bronchopleural fistula |
| Hospital C | PaO<sub>2</sub>/FiO<sub>2</sub> ratio <80 mmHg for more than 6 hours, PaO<sub>2</sub>/FiO<sub>2</sub> ratio <60 mmHg for more than 3 hours or bronchopleural fistula |
| Hospital D | Persistent PaO<sub>2</sub>/FiO<sub>2</sub> < 100 mmHg despite prone positioning |

FiO<sub>2</sub>: fraction of inspired oxygen; PaO<sub>2</sub>: partial pressure of oxygen.

At the moment of the analysis, 7646 patients were introduced in the registry, eight of those received ECMO support (0.1%). Patients were from four hospitals from two cities, with average 149 beds (ranging from 103 to 211 beds) and 39 intensive care unit beds(ranging from 30 to 51 beds). All of them were private hospitals and reference centers for COVID-19 treatment.

Controls were randomly selected from the same hospitals, among patients who had ARDS, needed to be treated at the intensive care unit and needed mechanical ventilation and pronation.

### Data collection

Demographic information, clinical characteristics, laboratory, imaging and outcome data were collected from medical records by trained hospital staff or interns, by using a prespecified case report form. Research Electronic Data Capture (REDCap) tools[17] hosted at the Telehealth Center, University Hospital, Universidade Federal de Minas Gerais. A detailed data management plan (DMP) was developed and provided to all participating centers. An online DMP training was mandatory before local research personnel could start collecting study data.

For the present analysis additional data of the ECMO procedure were collected: date of indication, date of the first session, whether it was veno-venous (VV-ECMO) or veno-arterial (VA-ECMO). Complications were already collected in the standardized form.

### Data quality assessment

We undertook comprehensive data quality checks to ensure high quality. Codes were developed in R software to identify outliers’ values likely related to data entry errors, based on expert-guided rules. Data was sent to each center for checking and correction. For the present analysis, as ECMO was a rare procedure, all cases were re-checked before the final analysis.

Transfers from one participant hospital to another were merged and considered as a single visit.

### Statistical analysis

To adjust for potential confounding variables, we used propensity score matching technique. Propensity scores were estimated by logistic regression, and the model included gender, age, number of comorbidities (hypertension, diabetes mellitus, obesity, coronary artery disease, heart failure, atrial fibrillation or flutter, cirrhosis, chronic obstructive pulmonary disease, cancer and previous stroke)[16], pronation, ARDS and hospital. Genetic matching method (MatchIt package in R software) was performed in a 5:1 ratio using generalized Mahalanobis distance with scaling factors chosen based on the smallest p-value in covariate balance tests.

Categorical data were presented as absolute numbers and proportions, and continuous variables were expressed as medians and interquartile ranges. The Fisher Exact test was used to compare the distribution of categorical variables, and the Wilcoxon-Mann–Whitney test for continuous variables. Results were considered statistically significant if the two-tailed P-value was < 0.05. All statistical analysis was performed with R software (version 4.0.2).

### Ethics

The study was approved by the National Commission for Research Ethics (CAAE 30350820.5.1001.0008). Individual informed consent was waived owing to the pandemic situation and the use of deidentified data, based on medical chart review only.

## Results

### Patient characteristics at hospital admission

From the 48 patients included in the study (Figure 1), eight received ECMO support during hospitalization, and 40 were matched controls. The median age of the entire sample was 60 (52.0-68.0) years-old and 87.5% were male. Hypertension (56.2%), diabetes mellitus (37.5%), obesity (22.9%), chronic heart failure (4.2%) were the most frequent comorbidities.

**Figure 1.**
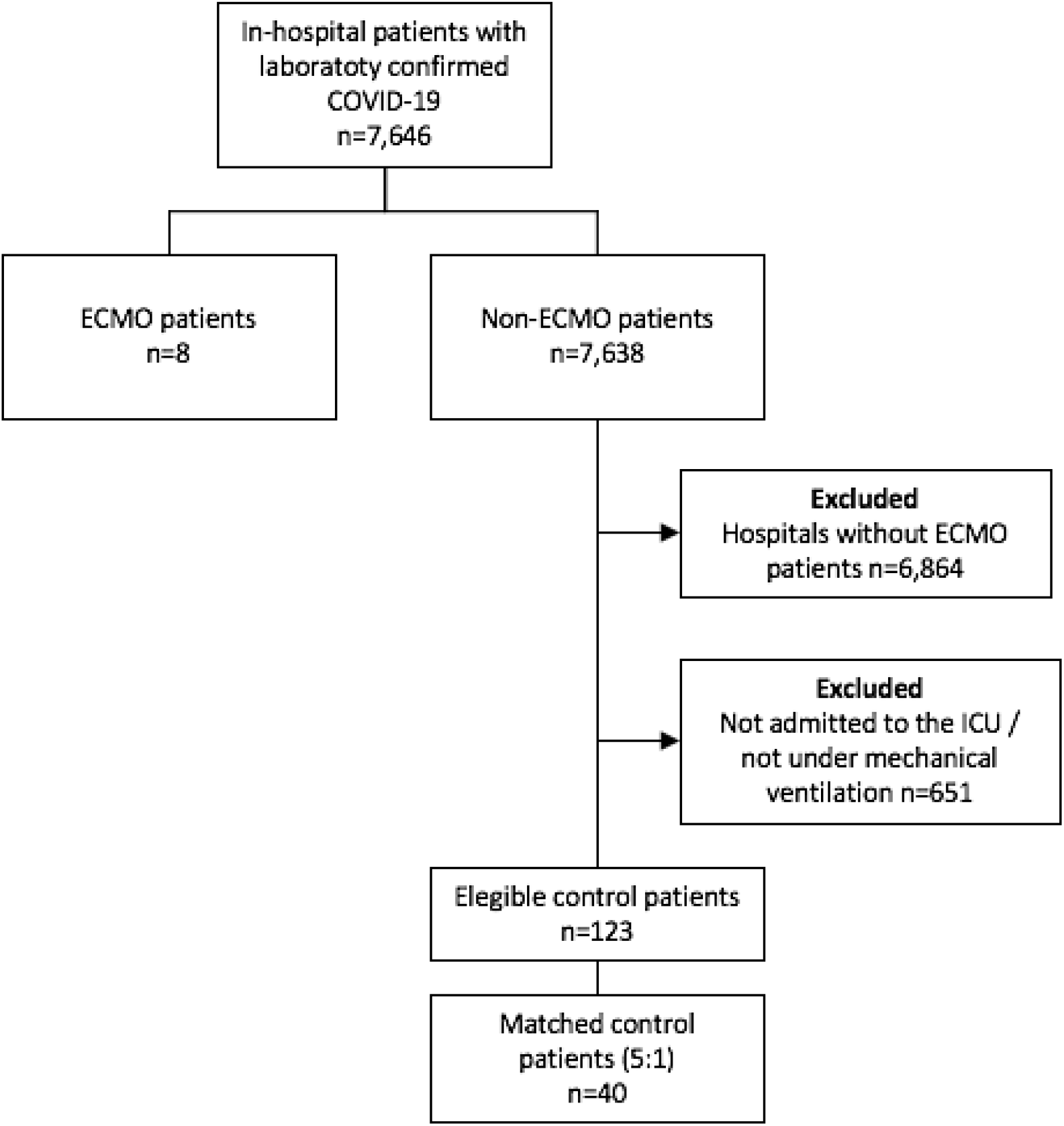
Flowchart of COVID-19 patients included in the study. ECMO: extracorporeal membrane oxygenation

When comparing patients who received ECMO support with controls, there were no significant differences in demographic characteristics and comorbidities (Table 2). Upon hospital admission, dyspnea, odynophagia and productive cough were present in more than one half of patients. The median time since from symptom onset to hospital admission was 6 (4.0-6.0) days. The two groups were similar in the clinical presentation (Table 3).

**Table 2.**
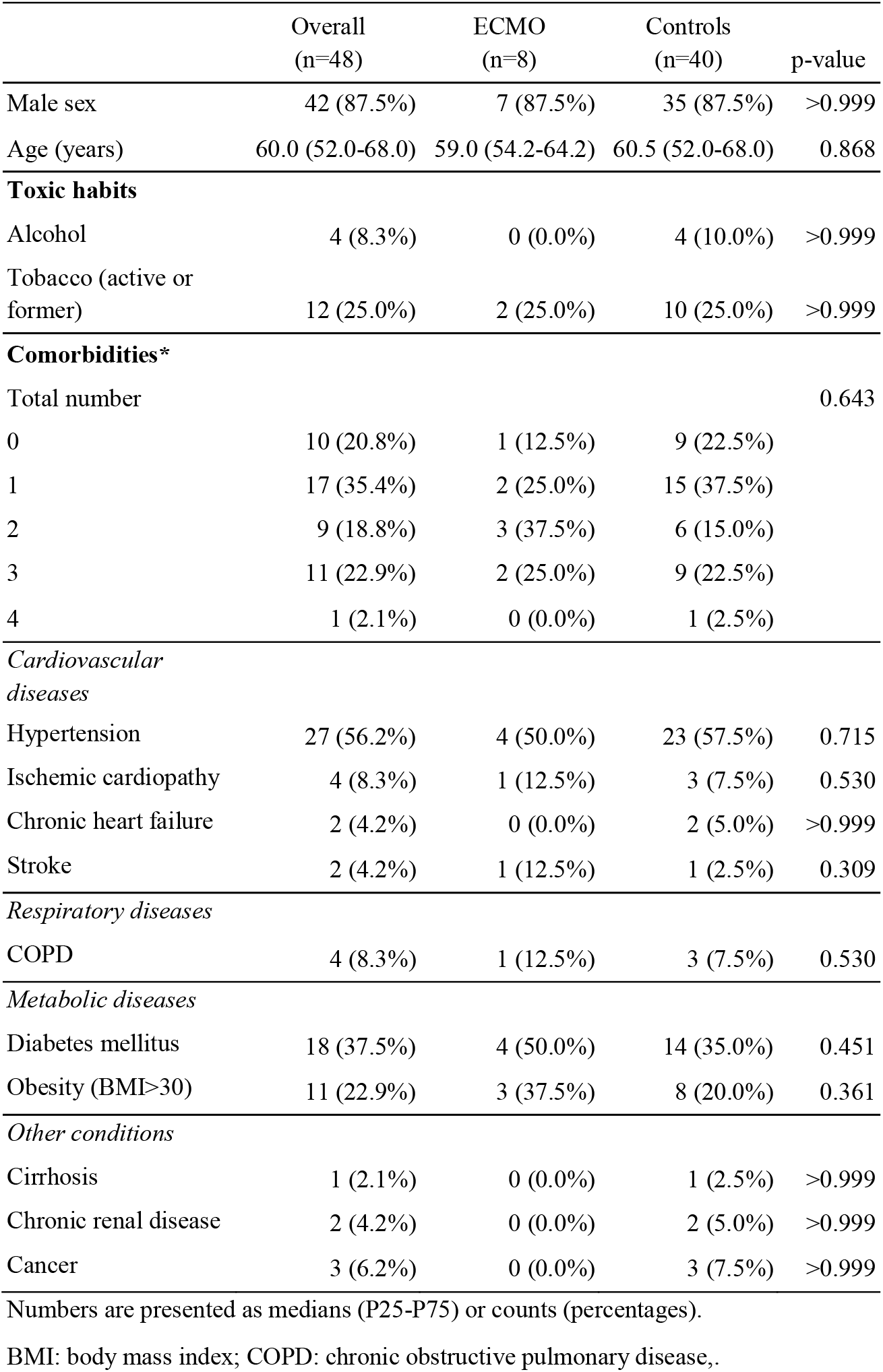
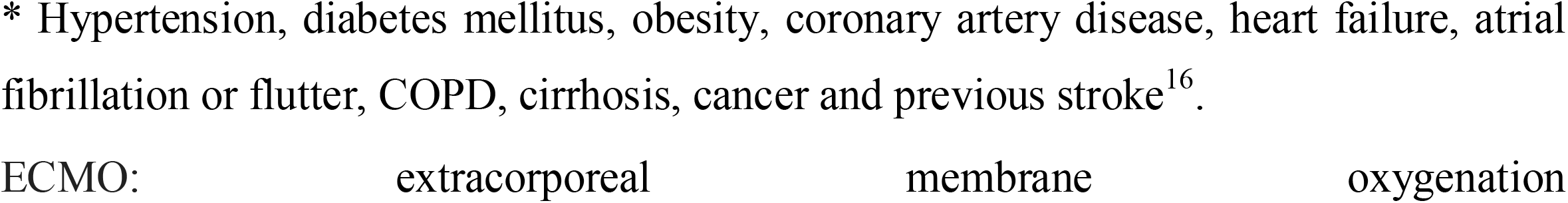
Demographic characteristics and medical history data at baseline.

**Table 3.** Clinical characteristics of the study population at baseline (n=48)

|  | Overall (n=48) |  | ECMO (n=8) |  | Control (n=40) |  |  |
| --- | --- | --- | --- | --- | --- | --- | --- |
|  | Frequency (%)<br>or median (IQR) | Non<br>missing<br>cases | Frequency (%)<br>or median (IQR) | Non<br>missing<br>cases | Frequency (%)<br>or median (IQR) | Non<br>missing<br>cases | P-value |
| **Symptoms** |  |  |  |  |  |  |  |
| Duration of symptoms (days) | 6.0 (4.0, 9.0) | 48 | 6.0 (4.5, 10.0) | 8 | 6.0 (4.0, 9.0) | 40 | 0.739 |
| Adynamic | 7 (14.6%) | 48 | 1 (12.5%) | 8 | 6 (15.0%) | 40 | >0.999 |
| Ageusia | 2 (4.2%) | 48 | 1 (12.5%) | 8 | 1 (2.5%) | 40 | 0.309 |
| Anosmia | 3 (6.2%) | 48 | 1 (12.5%) | 8 | 2 (5.0%) | 40 | 0.429 |
| Headache | 4 (8.3%) | 48 | 2 (25.0%) | 8 | 2 (5.0%) | 40 | 0.124 |
| Rhinorrhea | 8 (16.7%) | 48 | 2 (25.0%) | 8 | 6 (15.0%) | 40 | 0.605 |
| Diarrhea | 8 (16.7%) | 48 | 1 (12.5%) | 8 | 7 (17.5%) | 40 | >0.999 |
| Dyspnea | 28 (58.3%) | 48 | 3 (37.5%) | 8 | 25 (62.5%) | 40 | 0.251 |
| Fever | 25 (52.1%) | 48 | 5 (62.5%) | 8 | 20 (50.0%) | 40 | 0.703 |
| Hyporexia | 3 (6.2%) | 48 | 1 (12.5%) | 8 | 2 (5.0%) | 40 | 0.429 |
| Neurological manifestations | 0 (0.0%) | 48 | 0 (0.0%) | 8 | 0 (0.0%) | 40 |  |
| Myalgia | 10 (20.8%) | 48 | 3 (37.5%) | 8 | 7 (17.5%) | 40 | 0.336 |
| Nausea/vomiting | 8 (16.7%) | 48 | 3 (37.5%) | 8 | 5 (12.5%) | 40 | 0.116 |
| Productive cough | 4 (8.3%) | 48 | 1 (12.5%) | 8 | 3 (7.5%) | 40 | 0.530 |
| Dry cough | 26 (54.2%) | 48 | 5 (62.5%) | 8 | 21 (52.5%) | 40 | 0.710 |
| **Clinical assessment** |  |  |  |  |  |  |  |
| Glasgow <15 | 5 (10.4%) | 48 | 1 (12.5%) | 8 | 4 (10.0%) | 40 | >0.999 |
| Heart rate (HR) | 85.0 (76.5-103.0) | 47 | 85.0 (78.5- 113.2) | 8 | 85.0 (76.0-100.0) | 39 | 0.630 |
| HR ≥100 bpm | 14 (29.2%) | 48 | 3 (37.5%) | 8 | 11 (27.5%) | 40 | 0.676 |
| Respiratory rate (RR) | 20.0 (17.5, 24.0) | 35 | 18.0 (18.0, 18.0) | 5 | 20.0 (17.2, 25.5) | 30 | 0.366 |
| RR ≥24 mpm | 25 (52.1%) | 48 | 4 (50.0%) | 8 | 21 (52.5%) | 40 | >0.999 |
| SpO<sub>2</sub> | 93.0 (89.0, 94.2) | 48 | 92.0 (89.5, 92.2) | 8 | 93.5 (89.0, 95.0) | 40 | 0.249 |
| SpO<sub>2</sub><90% | 13 (27.1%) | 48 | 2 (25.0%) | 8 | 11 (27.5%) | 40 | >0.999 |
| Invasive ventilation | 4 (8.3%) | 48 | 1 (12.5%) | 8 | 3 (7.5%) | 40 | 0.530 |
| SBP ≤ 90mmHg | 2 (4.2%) | 48 | 0 (0.0%) | 8 | 2 (5.0%) | 40 | >0.999 |
| Inotropic drugs | 30 (62.5%) | 48 | 6 (75.0%) | 8 | 24 (60.0%) | 40 | 0.692 |
| SF ratio | 423.8 (154.3, 447.6) | 47 | 346.1 (138.2, 434.5) | 8 | 423.8 (168.3, 447.6) | 39 | 0.308 |

Numbers are presented as medians (P25-P75) or counts (percentages).
ECMO: extracorporeal membrane oxygenation; HR: heart rate; SpO<sub>2</sub>: peripheral oxygen saturation (%); SBP: systolic blood pressure; SF: SpO<sub>2</sub>/FiO<sub>2</sub>

There was no clinically relevant difference between the two groups concerning laboratory findings upon hospital admission (Table 4). With regards to the radiological findings, the low number of patients who underwent chest X-ray or CT-scan precluded any definitive comparison (Supplemental Table 1).

**Table 4.**
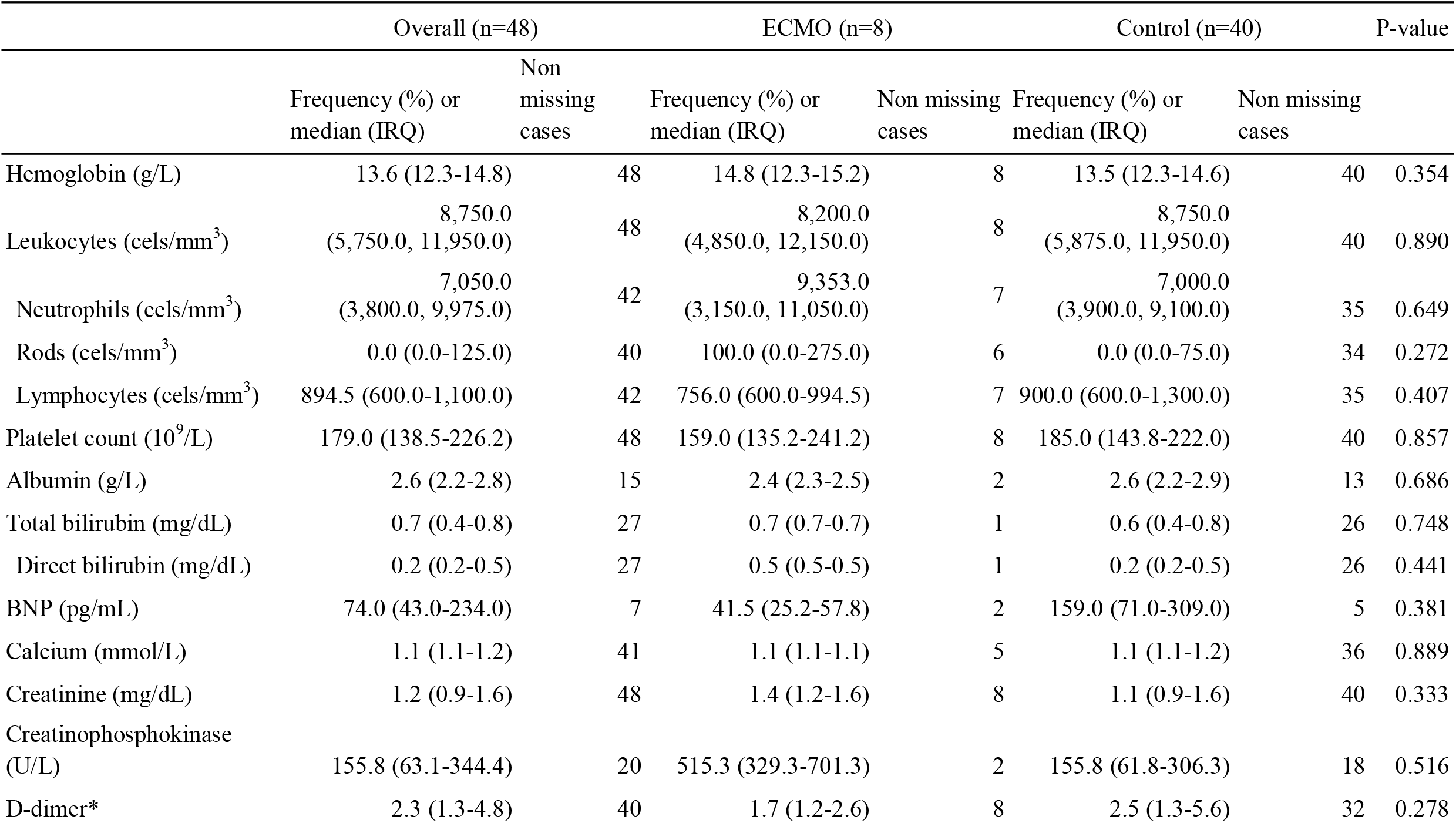
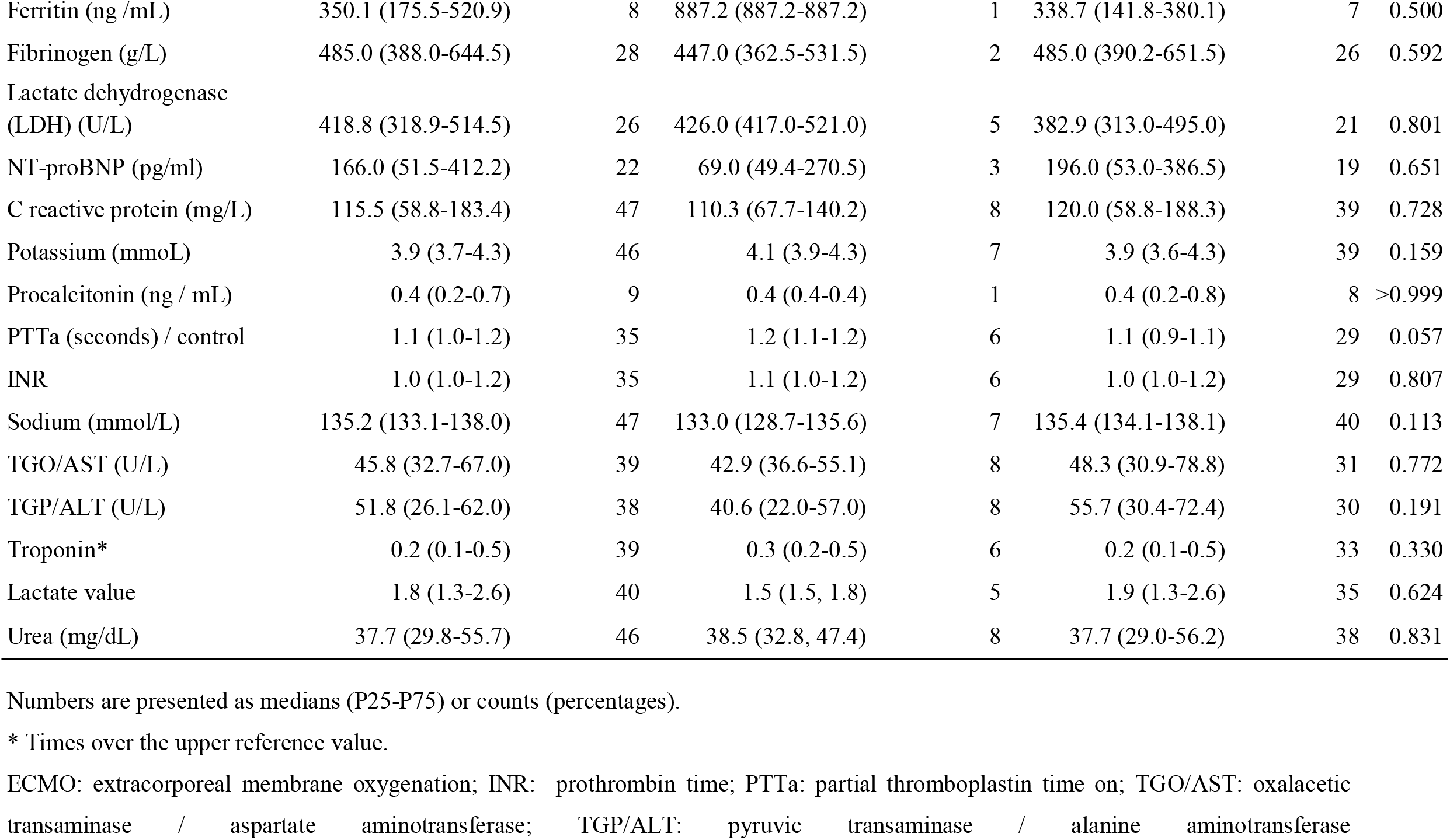
Laboratory parameters of the study population at baseline (n=48)

|  | Overall (n=48) |  | ECMO (n=8) |  | Control (n=40) |  | P-value |
| --- | --- | --- | --- | --- | --- | --- | --- |
|  | Frequency (%) or<br>median (IRQ) | Non<br>missing<br>cases | Frequency (%) or<br>median (IRQ) | Non missing<br>cases | Frequency (%) or<br>median (IRQ) | Non missing<br>cases |  |
| Hemoglobin (g/L) | 13.6 (12.3-14.8) | 48 | 14.8 (12.3-15.2) | 8 | 13.5 (12.3-14.6) | 40 | 0.354 |
| Leukocytes (cels/mm<sup>3</sup>) | 8,750.0<br>(5,750.0, 11,950.0) | 48 | 8,200.0<br>(4,850.0, 12,150.0) | 8 | 8,750.0<br>(5,875.0, 11,950.0) | 40 | 0.890 |
| Neutrophils (cels/mm<sup>3</sup>) | 7,050.0<br>(3,800.0, 9,975.0) | 42 | 9,353.0<br>(3,150.0, 11,050.0) | 7 | 7,000.0<br>(3,900.0, 9,100.0) | 35 | 0.649 |
| Rods (cels/mm<sup>3</sup>) | 0.0 (0.0-125.0) | 40 | 100.0 (0.0-275.0) | 6 | 0.0 (0.0-75.0) | 34 | 0.272 |
| Lymphocytes (cels/mm<sup>3</sup>) | 894.5 (600.0-1,100.0) | 42 | 756.0 (600.0-994.5) | 7 | 900.0 (600.0-1,300.0) | 35 | 0.407 |
| Platelet count (10<sup>9</sup>/L) | 179.0 (138.5-226.2) | 48 | 159.0 (135.2-241.2) | 8 | 185.0 (143.8-222.0) | 40 | 0.857 |
| Albumin (g/L) | 2.6 (2.2-2.8) | 15 | 2.4 (2.3-2.5) | 2 | 2.6 (2.2-2.9) | 13 | 0.686 |
| Total bilirubin (mg/dL) | 0.7 (0.4-0.8) | 27 | 0.7 (0.7-0.7) | 1 | 0.6 (0.4-0.8) | 26 | 0.748 |
| Direct bilirubin (mg/dL) | 0.2 (0.2-0.5) | 27 | 0.5 (0.5-0.5) | 1 | 0.2 (0.2-0.5) | 26 | 0.441 |
| BNP (pg/mL) | 74.0 (43.0-234.0) | 7 | 41.5 (25.2-57.8) | 2 | 159.0 (71.0-309.0) | 5 | 0.381 |
| Calcium (mmol/L) | 1.1 (1.1-1.2) | 41 | 1.1 (1.1-1.1) | 5 | 1.1 (1.1-1.2) | 36 | 0.889 |
| Creatinine (mg/dL) | 1.2 (0.9-1.6) | 48 | 1.4 (1.2-1.6) | 8 | 1.1 (0.9-1.6) | 40 | 0.333 |
| Creatinophosphokinase<br>(U/L) | 155.8 (63.1-344.4) | 20 | 515.3 (329.3-701.3) | 2 | 155.8 (61.8-306.3) | 18 | 0.516 |
| D-dimer* | 2.3 (1.3-4.8) | 40 | 1.7 (1.2-2.6) | 8 | 2.5 (1.3-5.6) | 32 | 0.278 |
| Ferritin (ng /mL) | 350.1 (175.5-520.9) | 8 | 887.2 (887.2-887.2) | 1 | 338.7 (141.8-380.1) | 7 | 0.500 |
| Fibrinogen (g/L) | 485.0 (388.0-644.5) | 28 | 447.0 (362.5-531.5) | 2 | 485.0 (390.2-651.5) | 26 | 0.592 |
| Lactate dehydrogenase (LDH) (U/L) | 418.8 (318.9-514.5) | 26 | 426.0 (417.0-521.0) | 5 | 382.9 (313.0-495.0) | 21 | 0.801 |
| NT-proBNP (pg/ml) | 166.0 (51.5-412.2) | 22 | 69.0 (49.4-270.5) | 3 | 196.0 (53.0-386.5) | 19 | 0.651 |
| C reactive protein (mg/L) | 115.5 (58.8-183.4) | 47 | 110.3 (67.7-140.2) | 8 | 120.0 (58.8-188.3) | 39 | 0.728 |
| Potassium (mmoL) | 3.9 (3.7-4.3) | 46 | 4.1 (3.9-4.3) | 7 | 3.9 (3.6-4.3) | 39 | 0.159 |
| Procalcitonin (ng / mL) | 0.4 (0.2-0.7) | 9 | 0.4 (0.4-0.4) | 1 | 0.4 (0.2-0.8) | 8 | >0.999 |
| PTTa (seconds) / control | 1.1 (1.0-1.2) | 35 | 1.2 (1.1-1.2) | 6 | 1.1 (0.9-1.1) | 29 | 0.057 |
| INR | 1.0 (1.0-1.2) | 35 | 1.1 (1.0-1.2) | 6 | 1.0 (1.0-1.2) | 29 | 0.807 |
| Sodium (mmol/L) | 135.2 (133.1-138.0) | 47 | 133.0 (128.7-135.6) | 7 | 135.4 (134.1-138.1) | 40 | 0.113 |
| TGO/AST (U/L) | 45.8 (32.7-67.0) | 39 | 42.9 (36.6-55.1) | 8 | 48.3 (30.9-78.8) | 31 | 0.772 |
| TGP/ALT (U/L) | 51.8 (26.1-62.0) | 38 | 40.6 (22.0-57.0) | 8 | 55.7 (30.4-72.4) | 30 | 0.191 |
| Troponin* | 0.2 (0.1-0.5) | 39 | 0.3 (0.2-0.5) | 6 | 0.2 (0.1-0.5) | 33 | 0.330 |
| Lactate value | 1.8 (1.3-2.6) | 40 | 1.5 (1.5, 1.8) | 5 | 1.9 (1.3-2.6) | 35 | 0.624 |
| Urea (mg/dL) | 37.7 (29.8-55.7) | 46 | 38.5 (32.8, 47.4) | 8 | 37.7 (29.0-56.2) | 38 | 0.831 |

Numbers are presented as medians (P25-P75) or counts (percentages).
\* Times over the upper reference value.
ECMO: extracorporeal membrane oxygenation; INR: prothrombin time; PTTa: partial thromboplastin time on; TGO/AST: oxalacetic transaminase / aspartate aminotransferase; TGP/ALT: pyruvic transaminase / alanine aminotransferase

There were no differences regarding the therapeutic strategy among both groups (Table 5), except for a trend of higher frequency of antiarrhythmic, antifungic, oseltamivir, statin (25.0% vs. 10.0%, p=0.258 for all of those) and hydroxycloroquine (37.5 vs. 15.0%, p=0.159) use in ECMO patients when compared to controls. No patient received remdesivir or tocilizumab.

**Table 5.**
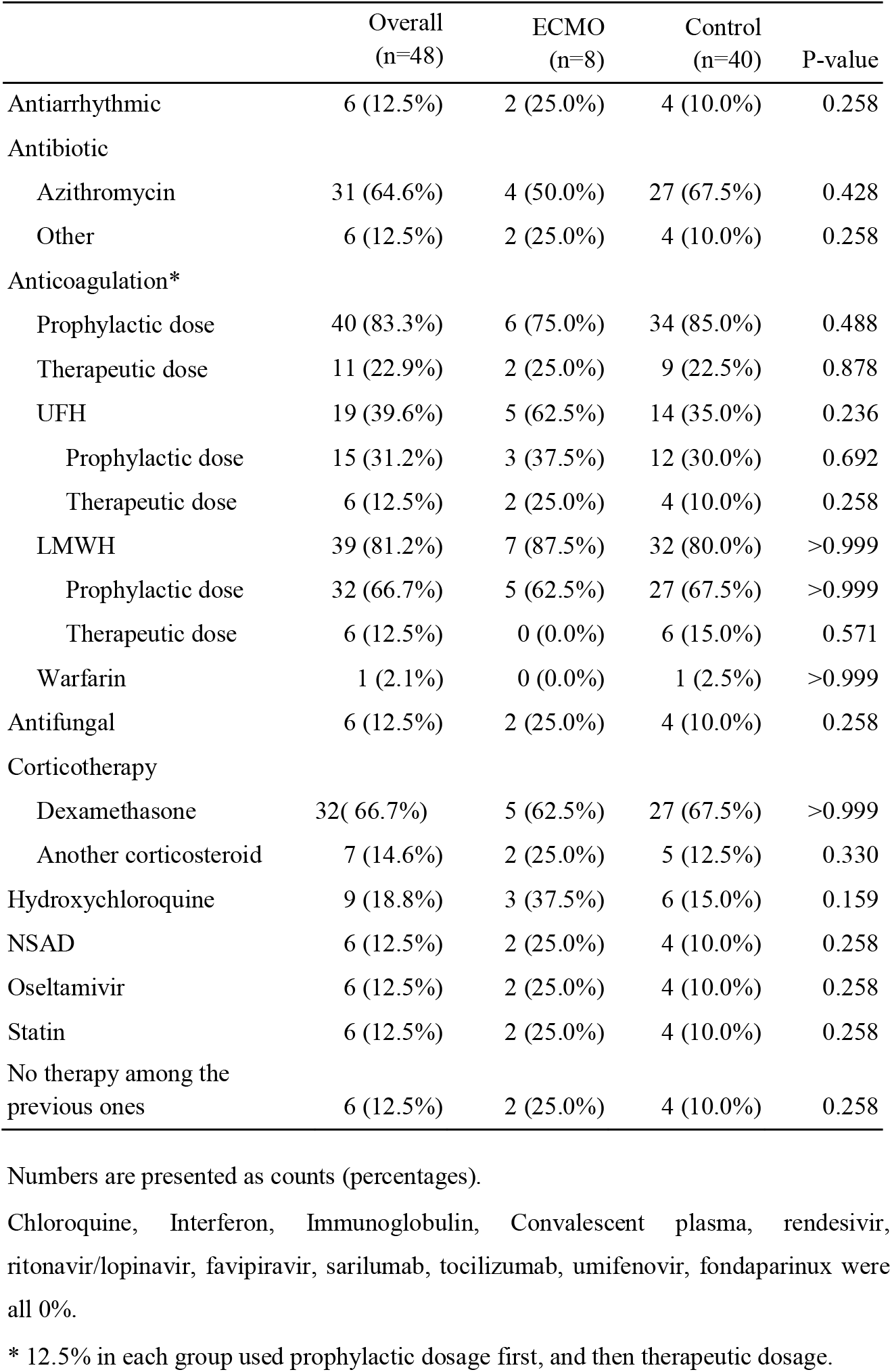
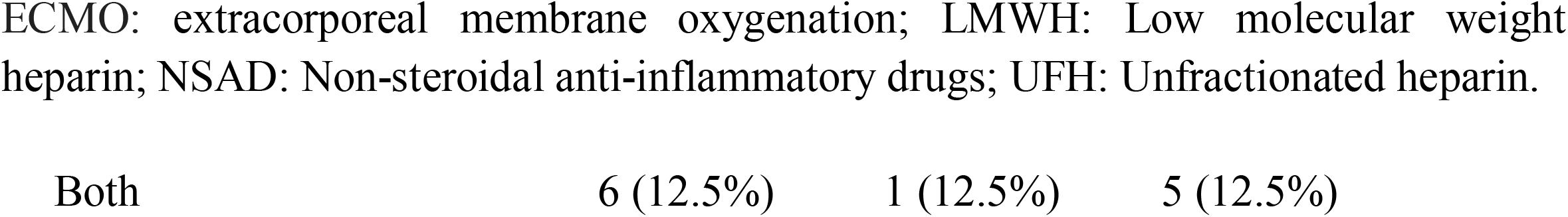
Medications of patients submitted to ECMO and controls (n=48)

|  | Overall<br>(n=48) | ECMO<br>(n=8) | Control<br>(n=40) | P-value |
| --- | --- | --- | --- | --- |
| Antiarrhythmic | 6 (12.5%) | 2 (25.0%) | 4 (10.0%) | 0.258 |
| Antibiotic |  |  |  |  |
| Azithromycin | 31 (64.6%) | 4 (50.0%) | 27 (67.5%) | 0.428 |
| Other | 6 (12.5%) | 2 (25.0%) | 4 (10.0%) | 0.258 |
| Anticoagulation* |  |  |  |  |
| Prophylactic dose | 40 (83.3%) | 6 (75.0%) | 34 (85.0%) | 0.488 |
| Therapeutic dose | 11 (22.9%) | 2 (25.0%) | 9 (22.5%) | 0.878 |
| UFH | 19 (39.6%) | 5 (62.5%) | 14 (35.0%) | 0.236 |
| Prophylactic dose | 15 (31.2%) | 3 (37.5%) | 12 (30.0%) | 0.692 |
| Therapeutic dose | 6 (12.5%) | 2 (25.0%) | 4 (10.0%) | 0.258 |
| LMWH | 39 (81.2%) | 7 (87.5%) | 32 (80.0%) | >0.999 |
| Prophylactic dose | 32 (66.7%) | 5 (62.5%) | 27 (67.5%) | >0.999 |
| Therapeutic dose | 6 (12.5%) | 0 (0.0%) | 6 (15.0%) | 0.571 |
| Warfarin | 1 (2.1%) | 0 (0.0%) | 1 (2.5%) | >0.999 |
| Antifungal | 6 (12.5%) | 2 (25.0%) | 4 (10.0%) | 0.258 |
| Corticotherapy |  |  |  |  |
| Dexamethasone | 32 (66.7%) | 5 (62.5%) | 27 (67.5%) | >0.999 |
| Another corticosteroid | 7 (14.6%) | 2 (25.0%) | 5 (12.5%) | 0.330 |
| Hydroxychloroquine | 9 (18.8%) | 3 (37.5%) | 6 (15.0%) | 0.159 |
| NSAD | 6 (12.5%) | 2 (25.0%) | 4 (10.0%) | 0.258 |
| Oseltamivir | 6 (12.5%) | 2 (25.0%) | 4 (10.0%) | 0.258 |
| Statin | 6 (12.5%) | 2 (25.0%) | 4 (10.0%) | 0.258 |
| No therapy among the previous ones | 6 (12.5%) | 2 (25.0%) | 4 (10.0%) | 0.258 |

|  |  |  |  |
| --- | --- | --- | --- |
| Both | 6 (12.5%) | 1 (12.5%) | 5 (12.5%) |

Overall, the median time from admission to ICU was 1.0 (0-3.0) day and the median hospital length of stay was 26.5 (IQR 15.5-44.2) days, and there was no difference between groups. The ECMO used in all eight patients was venovenous (V - V). The median time between stablishing criteria for ECMO and the initiation of the therapy was 0 (0 - 1) days.

Mortality was higher in ECMO patients (87.5 vs. 42.5%, p=0.048). There were no other significant differences in terms of clinical evolution and outcomes, despite a trend of a higher frequency of sepsis, disseminated intravascular coagulation and bleeding in the ECMO group (Table 6).

**Table 6.** Clinical evolution and outcomes of patients submitted to ECMO and controls (n=48)

|  | Overall (n=48) | ECMO (n=8) | Non-ECMO (n=40) | p-value |
| --- | --- | --- | --- | --- |
| Length of stay | 26.5 (15.5, 44.2) | 23.5 (19.0, 38.2) | 28.5 (13.5, 45.8) | 0.890 |
| Admission to ICU | 48 (100.0%) | 8 (100.0%) | 40 (100.0%) | 1.0 |
| Time from admission to ICU (days) | 1.0 (0.0, 3.0) | 1.0 (0.0, 2.2) | 1.5 (0.0, 3.0) | 0.657 |
| Days in ICU | 18.5 (10.8, 30.0) | 21.5 (18.5, 33.8) | 16.0 (8.8, 28.5) | 0.262 |
| Venous thromboembolic events | 7 (14.6%) | 1 (12.5%) | 6 (15.0%) | >0.999 |
| Mechanical ventilation | 48 (100.0%) | 8 (100.0%) | 40 (100.0%) | 1.0 |
| RRT | 22 (45.8%) | 6 (75.0%) | 16 (40.0%) | 0.119 |
| RRT before ECMO | NA | 4 (50.0%) | NA |  |
| Sepsis | 17 (35.4%) | 4 (50.0%) | 13 (32.5%) | 0.428 |
| Disseminated intravascular coagulation | 1 (2.1%) | 1 (12.5%) | 0 (0.0%) | 0.167 |
| Acute heart failure (new or chronic decompensated) | 2 (4.2%) | 0 (0.0%) | 2 (5.0%) | >0.999 |
| Acute heart failure (new onset) | 1 (2.1%) | 0 (0.0%) | 1 (2.5%) | >0.999 |
| Nosocomial infection | 5 (10.4%) | 1 (12.5%) | 4 (10.0%) | >0.999 |
| Bleeding | 2 (4.2%) | 1 (12.5%) | 1 (2.5%) | 0.309 |
| Death | 24 (50.0%) | 7 (87.5%) | 17 (42.5%) | 0.048 |

Numbers are presented counts (percentages).
ECMO: extracorporeal membrane oxygenation; ICU: intensive care unit; NA: not applicable; RRT: renal replacement therapy; VTE: venous thromboembolism.

ECMO related complications were observed in three (37.5%) patients: one patient (64 years-old, man) had severe bleeding in the cannulation site, kidney failure and need of renal replacement therapy, one (75 years-old, man) had massive thrombosis in the right atrium and the other (50 years-old, man) had kidney failure and need of renal replacement therapy.

## Discussion

This study describes a cohort of COVID 19 patients that received ECMO support in ICU from hospitals belonging to a large Brazilian COVID-19 Registry study. Overall, patients who received ECMO support had similar clinical characteristics, laboratory and outcomes to that patients did not receive ECMO support, matched by age, gender, number of comorbidities and hospital, except for a higher mortality and a trend of a higher frequency of complications, such as sepsis, disseminated intravascular coagulation and bleeding, in the ECMO group.

The use of ECMO as a rescue therapy in patients with severe ARDS secondary to viral infections has been established in the literature. In a cohort of patients with H1N1-related ARDS, Noah *et al*[18] demonstrated a hospital mortality of 23.7% for ECMO treated patients *vs* 52.5% for controls. Furthermore, in a retrospective study on MERS related ARDS, lower mortality was observed in the ECMO-treated cohort compared to controls (65 vs 100%, p=0.02)[3]. However, the EOLIA trial, the largest randomized controlled trial that evaluated the role of ECMO for ARDS, did not show a difference in 60-day mortality in the ECMO group when compared to the conventional management group (35 vs 46%, p=0.02)[4]. Two recent meta-analyses provided evidence that support use of ECMO for ARDS in adults[5, 6].

The pandemic has been placing significant stress on health care systems around the world, and hospitals were forced to prompt increase ICU capacity. Trained professionals suffered from work overload, burn out, or got sick, and in some instances inexperienced healthcare professionals had to assume positions. In this scenario, provision of ECMO may be challenging from both resource and ethical points of view[19]. The lack of full knowledge of the disease’s behavior, and the best way to approach it, the initial wait for a recovery that did not happen may have delayed the indication and the beginning of ECMO support in those patients, and this may be one of the reasons for our initial negative results, as well as the first series of literature. High mortality in the initial published experience[13] led some clinicians and investigators to recommend withholding ECMO support in patients with COVID-19[20]. Ruan *et al[7]* evaluated 137 COVID-19 patients and seven patients were treated with ECMO and there was 100% mortality despite ECMO use. Similar findings were reported by Zhou *et* al[9] and Yang *et al*[8], who observed mortality of 83 and 100%, respectively, in studies that included a very small number of patients.

It is well-known that COVID-19 mortality risk increases with increasing age and comorbidities[16, 21, 22]. In the present study, the small sample size precluded the assessment of specific risk factors for mortality in ECMO patients. Further studies are required to analyze which factors increase the mortality risk in those patients, in order to develop a proper patient care plan to try to reduce this high mortality.

It is not uncommon for patients on ECMO to develop acute kidney injury (AKI) within the first 48-72 h. The ECMO circuit can lead to an inflammatory reaction and subsequent AKI and renal replacement therapy simultaneously with ECMO can be necessary. Lastly, platelet consumption and potassium, magnesium, and phosphorous shifts have been observed in patients on ECMO and should be monitored and replaced accordingly[23].

The ELSO Registry provides data on 1035 ECMO - supported patients with COVID-19 who received care in 36 countries[12]. Estimated in-hospital mortality 90 days after ECMO initiation was 37.4% (95% CI 34·4–40·4), similar to previous mortality rates in non-COVID-19 ECMO-supported patients with ARDS and acute respiratory failure[4, 23], supporting the use of ECMO in COVID-19-related acute hypoxemic respiratory failure. However, these findings cannot be extrapolated to inexperienced centers. ELSO is clear in the recommendation against commissioning of new ECMO centers for the purposes of treating COVID-19 patients[24]. The training of the team, with ELSO certification, the multidisciplinary work, the 1: 1 patient per nurse ratio and the 2:1 specialist per patient ratio positively impact the results, and are essential requirements for the center to be accredited by the ELSO[24].

However, questions regarding its true role of ECMO remain unknown. The ELSO recommends to the accredited centers to preferentially offer ECMO to patients in whom outcomes are favorable (for example, those who are young, have a single organ failure, and were previously healthy)[24], differently to what was done in clinical practice, in this first phase of the pandemic.

Recently, Lebreton G. et al[25] presented the results of a well conduced study from the Extracorporeal Membrane Oxygenation Network, which involved 302 patients who underwent ECMO from 17 Greater Paris intensive care units, between March 8 and June 3, 2020. Before ECMO, 285 (94%) patients were prone positioned, median driving pressure was 18 cm H2O (14−21), and median ratio of the partial pressure of arterial oxygen to the fraction of inspired oxygen was 61 mm Hg (IQR 54−70). On ECMO, the most common adverse effects were a major bleeding event, renal replacement therapy and pulmonary embolism. Multiorgan failure were more frequently observed as causes of death 90 days after ECMO. At the end of 90 days, in general 138 (46%) patients were alive. Shorter time between intubation and ECMO, younger age (≤48 years), higher pre-ECMO renal component of the SOFA score, and treatment in centers managing at least 30 venovenous ECMO cases annually were independently associated with improved 90-day survival (60%), among ECMO-assisted patients with COVID-19 center’s experience in venovenous ECMO during the previous year was a strongly predictor of survival. Therefore, organization and experience are fundamental to best results in patients with COVID-19 and severe ARDS requiring ECMO.

Other main factors that should be taken into consideration are its feasibility and ethical dilemmas. In most parts of the world, patients may not be able to benefit from this modality of treatment due to the lack of availability and its high cost. In this

Brazilian Registry which included 37 centers, only 0.1% of patients from four hospitals were submitted to ECMO support.

The high cost associated with the ECMO use also may be considered as a critical factor in the decision about using this technology and makes its inappropriate use a high waste for the system (or families in the specific case from this Brazilian cohort)[26]. In times when healthcare labor resources are scarce, it is of utmost importance to consider that the elevated cost of using ECMO is not only associated with the technology price, but much more in the demand for nursing and cardiology assistants and for ICU beds that those patients require[27]. In a Dutch study, the mean cost of ECMO patients was (Euros) 106.263 and 52% of these costs arose from nursing care and only 11% from the ECMO surgery and materials[28]. In Brazil, a short cohort study in 2018 demonstrated that ECMO materials and surgery represented 11% of the cost per patient for surviving patients, 30% for those who died[29]. These previous experiences highlight the importance of taking the right decision on using the technology, at the right time.

Additionally, it is of utmost importance to consider all costs, not only the cost and the right use of the equipment and the timely indication for the procedure, but also the suitable hospital structure and trained human resources required for the best use of the technology [26].

In the present study, all ECMO patients were submitted to veno-venous ECMO, what is in line with a recent systematic review, in which the majority of COVID-19 patients who require ECMO were submitted to this modality (73%), while veno-arterial ECMO were performed in a minority proportion of patients (3.3%), 2.5% moved between these two types or needed a more specific ECMO according to the disease prognosis, and in 20.8% of patients the type was not mentioned.

This study has limitations. First, it was not a randomised controlled trial and thus we cannot draw any definitive conclusions as to whether ECMO should be used in patients with COVID-19 and severe respiratory failure. However, our results are consistent with previously reported survival rates in acute hypoxaemic respiratory failure, supporting current ELSO recommendations that centres experienced in ECMO should withhold its use in refractory COVID-19-related respiratory failure in situations of healthcare system collapse. The ECMO may be demanding from both resource and ethical points. In a crisis scenario, the allocation of resources, whether human or financial, is challenging. Stricter selection criteria are recommended in order to use this resource for patients with greater chances of recovery. ECMO is recommended for the largest and certified centers only and it is not recommended when the system is overwhelmed[24].

A lot of research is still needed to show the role of ECMO support in COVID19 patients’ survival. In Brazil, in the second phase of the pandemic those centers may have improved ECMO procedures, as they established protocols and tried to avoid delayed indication, when it might have been too late to be of any benefit. We therefore hypothesize that future analyzes of data from a second phase of the pandemic might provide more encouraging mortality outcomes than those found in the first phase of lesser expertise with ECMO in patients with COVID-19 and even less knowledge of the disease in general.

The inclusion of ECMO as a procedure sponsored by the public health system is presently being assessed by the Brazilian National Commission for the Incorporation of Health Technology (CONITEC), but we believe that careful and adequate assessment of clinical practice outcomes is of utmost importance before ECMO is incorporated for the health system. Further analysis of the COVID-19 Brazilian Registry may provide interesting results of the comparison of different phases of the pandemic.

## Data Availability

Data are available upon reasonable request.

## Acknowledgments

We would like to thank the hospitals which are part of this collaboration, for supporting this project. We also thank all the clinical staff at those hospitals, who cared for the patients, and all undergraduate students who helped with data collection.

## Data availability statement

Data are available upon reasonable request.

---

**Supplemental Table 1.** Radiological characteristics of the study population at baseline.

|  | Overall<br>(n=48) | ECMO<br>(n=8) | Control<br>(n=40) | P-value |
| --- | --- | --- | --- | --- |
| **X-ray at admission (24h)** | **31 (64.6%)** | **3 (37.5%)** | **28 (70.0%)** | **0.112** |
| Atelectasis | 0 (0.0%) | 0 (0.0%) | 0 (0.0%) |  |
| Consolidation | 0 (0.0%) | 0 (0.0%) | 0 (0.0%) |  |
| Diffuse interstitial infiltrate | 1 (3.2%) | 0 (0.0%) | 1 (3.6%) | >0.999 |
| Focal interstitial infiltrate | 0 (0.0%) | 0 (0.0%) | 0 (0.0%) |  |
| Bilateral ground-glass opacity | 18 (58.1%) | 0 (0.0%) | 18 (64.3%) | 0.064 |
| Single-sided frosted glass opacity | 0 (0.0%) | 0 (0.0%) | 0 (0.0%) |  |
| None | 12 (38.7%) | 3 (100.0%) | 9 (32.1%) | 0.049 |
| **X-ray Follow up** | **45 (93.8%)** | **7 (87.5%)** | **38 (95.0%)** | **0.429** |
| Atelectasis | 1 (2.2%) | 0 (0.0%) | 1 (2.6%) | >0.999 |
| Consolidation | 5 (11.1%) | 1 (14.3%) | 4 (10.5%) | >0.999 |
| Diffuse interstitial infiltrate | 3 (6.7%) | 1 (14.3%) | 2 (5.3%) | 0.405 |
| Focal interstitial infiltrate | 0 (0.0%) | 0 (0.0%) | 0 (0.0%) |  |
| Bilateral ground-glass opacity | 36 (80.0%) | 5 (71.4%) | 31 (81.6%) | 0.614 |
| Single-sided ground-glass | 2 (4.4%) | 0 (0.0%) | 2 (5.3%) | >0.999 |
| None of the above | 7 (15.6%) | 1 (14.3%) | 6 (15.8%) | 1 |
| Progression | 8 (17.8%) | 1 (14.3%) | 7 (18.4%) | >0.999 |
| **Chest CT admission** |  |  |  |  |
| Atelectasis | 0 (0.0%) | 0 (0.0%) | 0 (0.0%) |  |
| Consolidation | 4 (18.2%) | 0 (0.0%) | 4 (22.2%) | 0.554 |
| Pleural effusion | 3 (13.6%) | 0 (0.0%) | 3 (16.7%) | >0.999 |
| Bilateral ground-glass pattern | 18 (81.8%) | 3 (75.0%) | 15 (83.3%) | >0.999 |
| Peripheral ground-glass pattern | 5 (22.7%) | 1 (25.0%) | 4 (22.2%) | >0.999 |
| Mosaic paving | 3 (13.6%) | 1 (25.0%) | 2 (11.1%) | 0.470 |
| None | 0 (0.0%) | 0 (0.0%) | 0 (0.0%) |  |
| **Chest CT follow up** | **11 (22.9%)** | **1 (12.5%)** | **10 (25.0%)** | **0.661** |
| Atelectasis | 2 (18.2%) | 0 (0.0%) | 2 (20.0%) | >0.999 |
| Consolidation | 3 (27.3%) | 0 (0.0%) | 3 (30.0%) | >0.999 |
| Pleural effusion | 3 (27.3%) | 0 (0.0%) | 3 (30.0%) | >0.999 |
| One-sided ground-glass pattern | 1 (9.1%) | 0 (0.0%) | 1 (10.0%) | >0.999 |
| Bilateral ground-glass pattern | 8 (72.7%) | 1 (100.0%) | 7 (70.0%) | >0.999 |
| Peripheral ground-glass pattern | 3 (27.3%) | 0 (0.0%) | 3 (30.0%) | >0.999 |
| Mosaic paving | 2 (18.2%) | 0 (0.0%) | 2 (20.0%) | >0.999 |
| None | 0 (0.0%) | 0 (0.0%) | 0 (0.0%) |  |
| Progression | 11 (100.0%) | 1 (100.0%) | 10 (100.0%) | 1 |

Numbers are presented counts (percentages).
CT: computerized tomography; ECMO: extracorporeal membrane oxygenation.

